# Sulodexide in the treatment of patients with early stages of COVID-19: a randomised controlled trial

**DOI:** 10.1101/2020.12.04.20242073

**Authors:** Alejandro J. Gonzalez-Ochoa, Joseph D. Raffetto, Ana G. Hernández, Nestor Zavala, Obed Gutiérrez, Arturo Vargas, Jorge Loustaunau

## Abstract

**Background:** Targeting endothelial cells has been suggested for the treatment of patients with COVID-19 and sulodexide has pleiotropic properties within the vascular endothelium that can prove beneficial to the same. We aimed to evaluate the effect of sulodexide when used in the early clinical stages of COVID-19.

**Methods:** We conducted a single-centre, outpatient setting, randomised controlled trial with a parallel-group design in Mexico. Including patients within three days of clinical symptom onset, who were at a high risk of severe clinical progression due to chronic comorbidities. Participants were randomly allocated to receive an oral dose of sulodexide (500 LRU twice a day) or the placebo for 21 days. Primary outcomes were need and length of hospitalisation, need and length of oxygen support.

**Results:** Between June 5 and August 30, 2020, 243 patients were included in the “per-protocol” analysis. One hundred twenty-four of them received sulodexide, while 119 received placeboes. At 21 days follow-up, 22 of 124 patients required hospitalisation in the sulodexide group compared to 35 of 119 in the placebo group [relative risk (RR), 0·6; 95% confidence interval (CI), 0·37-0·96; p=0·03]. Fewer patients required oxygen support in the sulodexide group [37 of 124 vs. 50 of 119; RR, 0·71; 95% CI, 0·5 to 1; p=0·05], and for fewer days (9±7·2 in the sulodexide group vs. 11·5±9·6 in the placebo group; p=0·02). There was no between-group difference concerning the length of hospital stay.

**Interpretation:** Early intervention in COVID-19 patients with sulodexide reduced hospital admissions and oxygen support requirements, although with no significant effect on mortality. This has beneficial implications in the patient well-being, making sulodexide a favourable medication until an effective vaccine or an antiviral becomes available.

**Funding:** Researcher independently initiated, partially funded by Alfasigma, Mexico.

Listed in the ISRCTN registry under ID ISRCTN59048638.

## Introduction

The emergence of the novel coronavirus disease 2019 (COVID-19) developed into a pandemic that changed our way of life to a degree yet to be determined. The reported percentage of infected patients who need hospital care is between 15-25%. However, its high rate of contagiousness has maimed healthcare systems worldwide due to the overall load of patients in need of hospital care. The reduction in space available for other patients in need of critical care has further aggravated an already depleted healthcare infrastructure, especially in low socioeconomic countries^1-3^. Early intervention to prevent the more aggressive forms of COVID-19 could significantly impact hospital resources, aiding the resumption of standard care for individuals with non-COVID-19 medical conditions.

Severe acute respiratory syndrome coronavirus 2 (SARS-CoV-2) infects the host via the angiotensin-converting enzyme 2 (ACE2) receptor, which is expressed in several organs apart from the lungs— most importantly the heart, kidney and the vascular endothelium of both small and large arteries as well as veins^4,5^. The vascular endothelium is an active paracrine, endocrine and autocrine organ that is indispensable in the regulation of vascular tone and the maintenance of vascular homeostasis. The endothelial surface layer on the lungs plays a critical role in the host immune response to the virus, both as an effector and as a target organ. There is evidence of endothelial viral inclusion^6^ and diffuse endothelial inflammation (endothelialitis), triggering a systemic release of inflammatory cytokines. This response produces an imbalance between the excessive formation of reactive oxygen species (ROS) and the antioxidant defence capacity, which is considered a hallmark of endothelial dysfunction. Under such conditions, the endothelium’s protective properties are diminished, resulting in a proinflammatory and prothrombotic state that is worsened due to blood flow stasis through reactive vasoconstriction. Virchow’s triad of events could explain the systemic impaired function in different vascular beds and their clinical sequelae in some patients^7-9^. COVID-19-endothelialitis could be particularly relevant for vulnerable patients with pre-existing endothelial dysfunction, which is associated with male sex, older age and chronic comorbidities— all of which have been linked to adverse outcomes^10,11^. To date, pulmonary endothelial cells (ECs) have been largely overlooked as a therapeutic target in COVID-19^12^.

Sulodexide is composed of two glycosaminoglycans (GAGs), namely a fast-moving heparin fraction (80%) and dermatan sulphate (20%), each with several pleiotropic endothelial actions that can be beneficial in COVID-19 patients^13^. As a precursor for the synthesis of GAGs, it can help restore shredded endothelial glycocalyx and prevent further degradation^14,15^. The improvement of the glycocalyx integrity not only restores the endothelium barrier function, it also allows the endothelium to better modulate the generation of key inflammatory molecules, such as IL1β, IL6, IL8 and TNFα, while also downregulating its response to them^16,17^. The heparin compound adds an antithrombotic and profibrinolytic effect that may be important against the procoagulant state caused by SAR-CoV-2; moreover, it may exert added anti-inflammatory effects^18,19^.

We hypothesised that sulodexide’s pleiotropic properties could provide an antithrombotic effect, improve endothelial integrity and decrease inflammatory responses that would limit the damage caused by SARS-COV-2, thereby improving the clinical outcome. With this premise, we developed a randomised trial to evaluate whether sulodexide has a positive effect in mitigating the severe clinical progression rate of the disease, culminating in decreased hospital admission, decreased oxygen support use and decreased mortality.

## Methods

### Study design

This is a prospective, randomised placebo-controlled trial with a parallel-group design evaluating the clinical outcome of consecutive patients who suffer early clinical stages of COVID-19. This was defined as any two symptoms, including headache, fever or cough, accompanied by one of the following: diarrhoea, body/muscle ache, loss of smell/taste, difficulty breathing, conjunctivitis, runny nose or chest pain^20^, within three days of onset or less.

The recruiting period extended from June 5 to August 5, 2020. The location of the study site is in the city of San Luis R.C., Sonora, a border port of entry in the northwest region of Mexico— with a reported total of 28,990 confirmed COVID-19 cases as of August 5, 2020— at a 100 miles radius, including cities on the USA side of the border. We conducted this trial according to the Declaration of Helsinki, and it was board reviewed by the Universidad Autonoma de Baja California Faculty of Medicine Mexicali Ethics and Investigation Committee, being designated with approval number FMM/CEI/0011/2020-2.

### Patients

Anticipating difficulties in recruiting eligible patients, we utilised social media outreach and contacted primary care physicians in government as well as private practices for early referral. We reached out to healthcare workers at COVID-19 converted hospitals and household members of known positive COVID-19 patients, owing to their high risk of infection and first-hand knowledge of symptoms.

Virtual communication was implemented for screening patient eligibility. Inclusion criteria were as follows: male or female, age over 40 years, with suspected COVID-19 clinical symptom within 3 days of onset or less and high risk (>50%) of a severe clinical disease progression according to the percentage of risk given by the COVID-19 Health Complication (C19HC) calculator (IMSS, Gobierno de Mexico),^21^ which considers different chronic comorbidities described in *table 1*. Important exclusion criteria included: a negative COVID-19 test, already in hospital care, prolonged anticoagulation, or venous thromboembolism in the past six months (*table 2*). The initiation of anticoagulant medication at prophylactic dose while the trial was ongoing was not a criteria for elimination, although a stricter follow-up was implemented due the possible risk of bleeding complications.

**Table 1.**
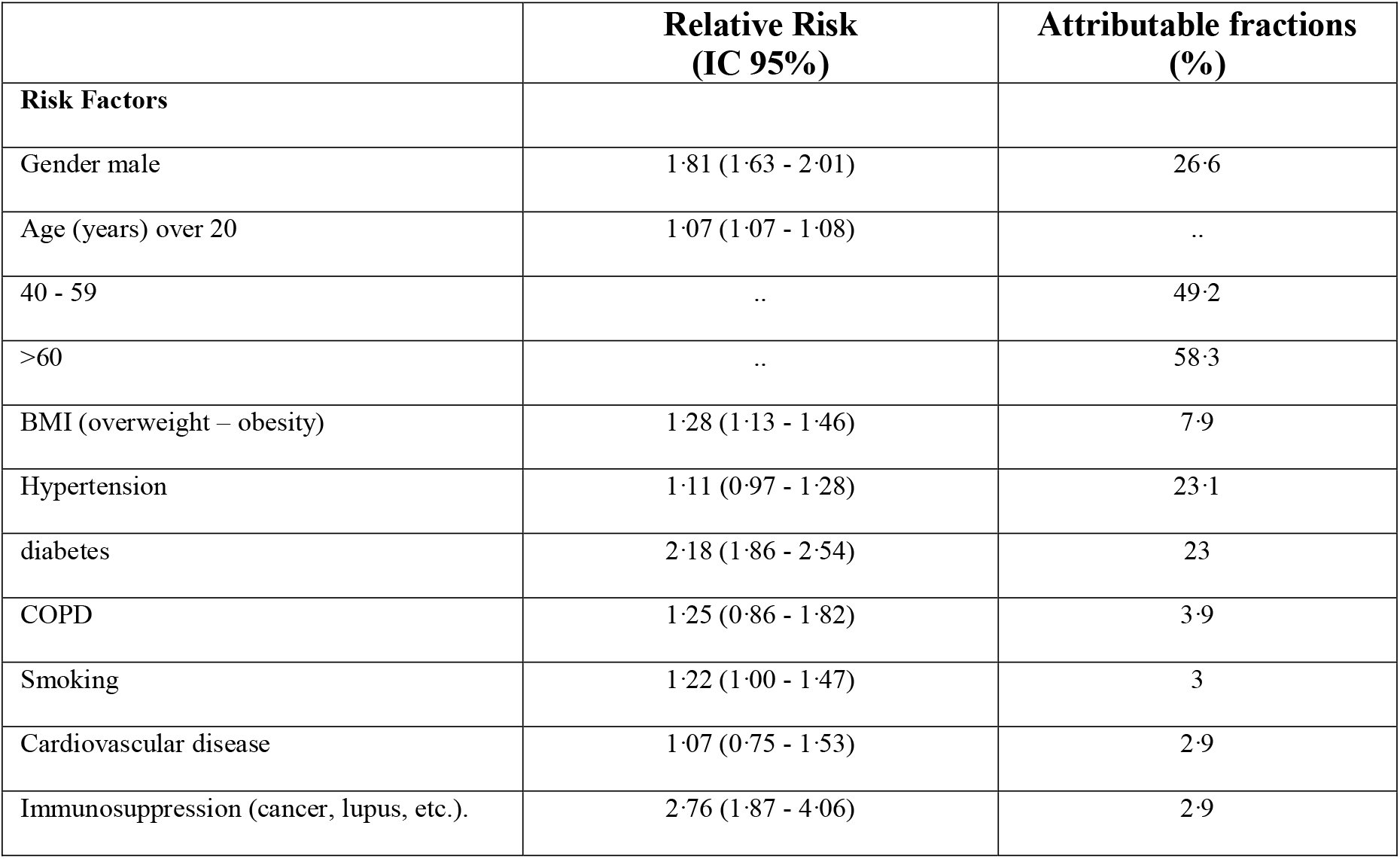
COVID-19 Health Complication (C19HC) calculator. The calculator is available online. An automatic risk calculation result is given once inputted the requested data. The calculation is obtained according to the unconditional multiple regression model, through the algorithm sum of the weight given by each risk factor relative risk and their attributable fractions. A result of <50% = medium risk, 50-80% = high risk, and >80% = very high risk. IC = confidence interval BMI= body-mass index is the weight in kilograms divided by the square of the height in meters, were <18·5 = underweight, 18·5-24·9 = normal, 25-29·9 = overweight, >30 = obesity. COPD= Chronic obstructive pulmonary disease

**Table 2.**
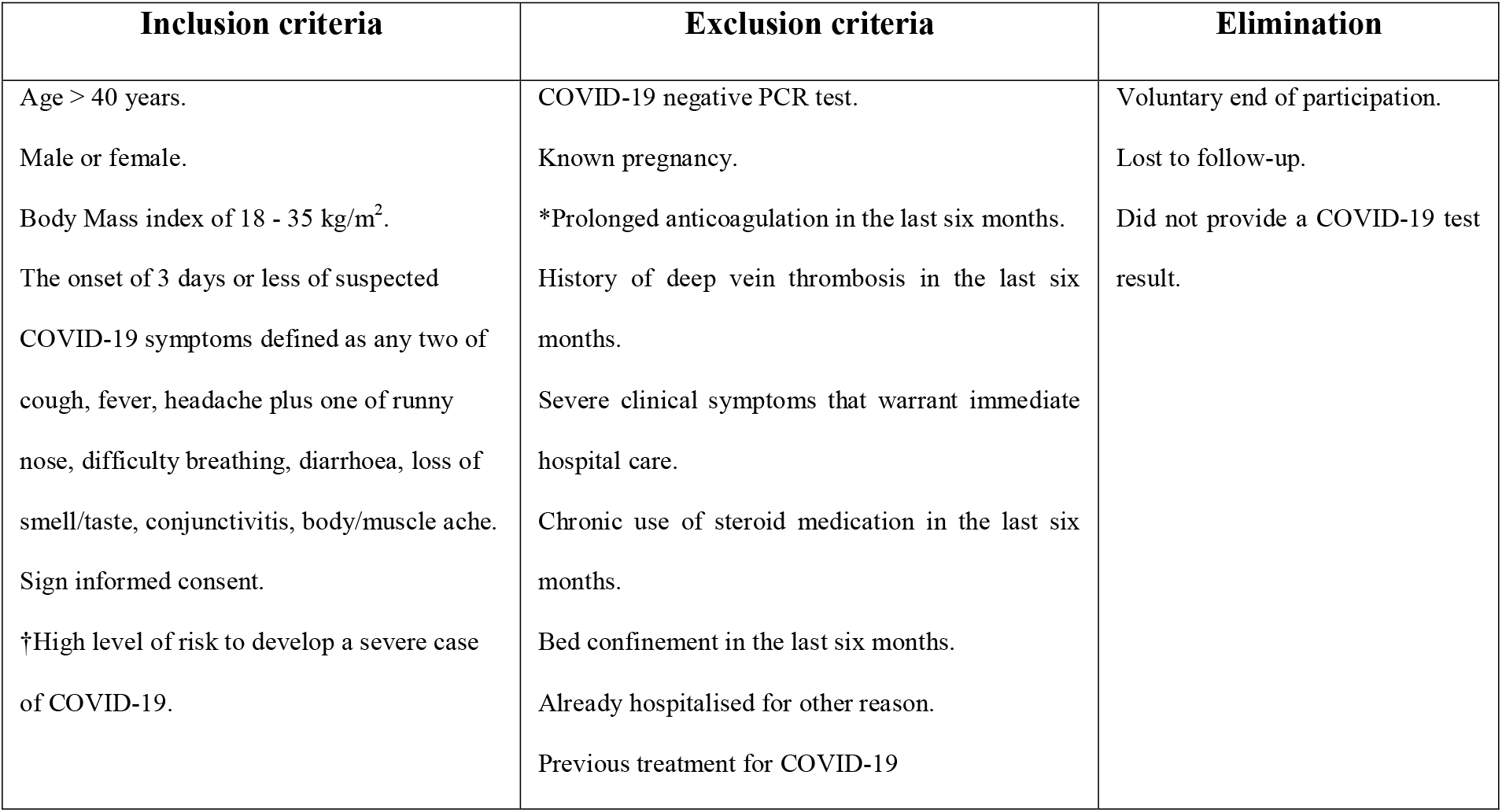
Inclusion, exclusion, and elimination criteria. †According to the COVID-19 Health Complication calculator (IMSS, Gobierno de Mexico). BMI = body mass index. * Start of anticoagulation after trial inclusion was not a criterion for exclusion.

If eligible, an informed consent form was signed, and the patient was scheduled at the earliest for blood exams and the COVID-19 polymerase chain reaction (PCR) test.

### Intervention

Sequential randomisation for group allocation was performed at the research site with the computer software aid provided by Castor Electronic Data Capture, Amsterdam, The Netherlands). The software generated a permuted block randomisation sequence in a 1:1 ratio and under no strata. Only the lead researcher knew the allocation result; neither the rest of the collaborators nor the patients had any knowledge of the same.

The research site dispersed 250RLU capsules of masked sulodexide (Vessel due F, Alfasigma Mexico) or masked placebo following a seven-day supply schedule. The indicated dose was 500RLU twice daily for a duration of 3 weeks. Although 250RLU twice a day has demonstrated effective plasma concentrations *in vitro* and is regularly prescribed in daily practice^13^, we chose the 500RLU dosing regimen based on the SURVET study, in which an antithrombotic effect was safely achieved in a clinical setting^22^. We prescribed placebo tablets for the control group based on an identical regimen. As a result of local logistic limitations imposed due to the regional pandemic lockdown, as well as to verify timely medication dispensation, the lead researcher was not blinded to group allocation. Other than the study medication and the follow-up related to the study endpoints, the research site was not involved in the disease’s primary treatment, as a means to prevent bias. Patients were encouraged to continue with the standard of care provided by their healthcare providers^23^. Although some of the study collaborators outside of the research site could be involved in the patient complementary treatment, they were blinded to group allocation.

Different authorised laboratory sites oversaw the confirmatory COVID-19 PCR test. Since the result could take several days to become available, the participant continued the follow-up as scheduled. If confirmed positive, the participant would continue in the trial; otherwise, if negative, we suspended the medication and excluded the patient from data analysis.

We performed the study endpoint follow-up via virtual communication with participants or household members, every seven days or as deemed necessary, during the 3-week participation period. If no virtual form of contact was possible, we scheduled a field visit to the participant’s home. At day 14 of follow-up, a new blood sample test was scheduled for secondary endpoints evaluation. Strict safety protocols by the laboratory staff members were followed. If we failed to contact the participant during the follow-up period, and no data were available other than the initial inclusion survey, the patient was excluded from the final analysis after eliminating mortality as the cause of the inability to follow up.

If the patient’s symptoms worsened, we instructed a hospital visit with the corresponding patient healthcare provider. A hospital visit to the emergency department was not reported as a study endpoint unless it resulted in formal admission to the COVID-19 hospital ward. Hospitals posted in-house protocols for clinical management and hospital admission— including, but not limited to, respiratory failure (oxygen saturation <90%, severe hypoxemia [partial pressure of oxygen <60 mm Hg] or breathing rate >30 breaths per min, while breathing ambient air); abnormal chest X-ray compatible with COVID-19-associated pneumonia; and relevant clinical alterations, including hemodynamic, hepatic, renal or haematological derangements, along with clinically significant laboratory abnormalities^24^.The indication for hospital admission or need for at-home oxygen support was left to the discretion of the emergency department physician in charge, who was blinded to group allocation. If the patient required hospital care, we suspended the oral dose of sulodexide or placebo. Although the research team was not involved in any treatment decision during hospital care, follow-up continued past the stipulated 3-week period until we could define an outcome, or until the trial time ended. After hospital discharge, sulodexide was not re-initiated.

The CONSORT diagram flow is detailed in *figure 1*.

**Figure 1.**
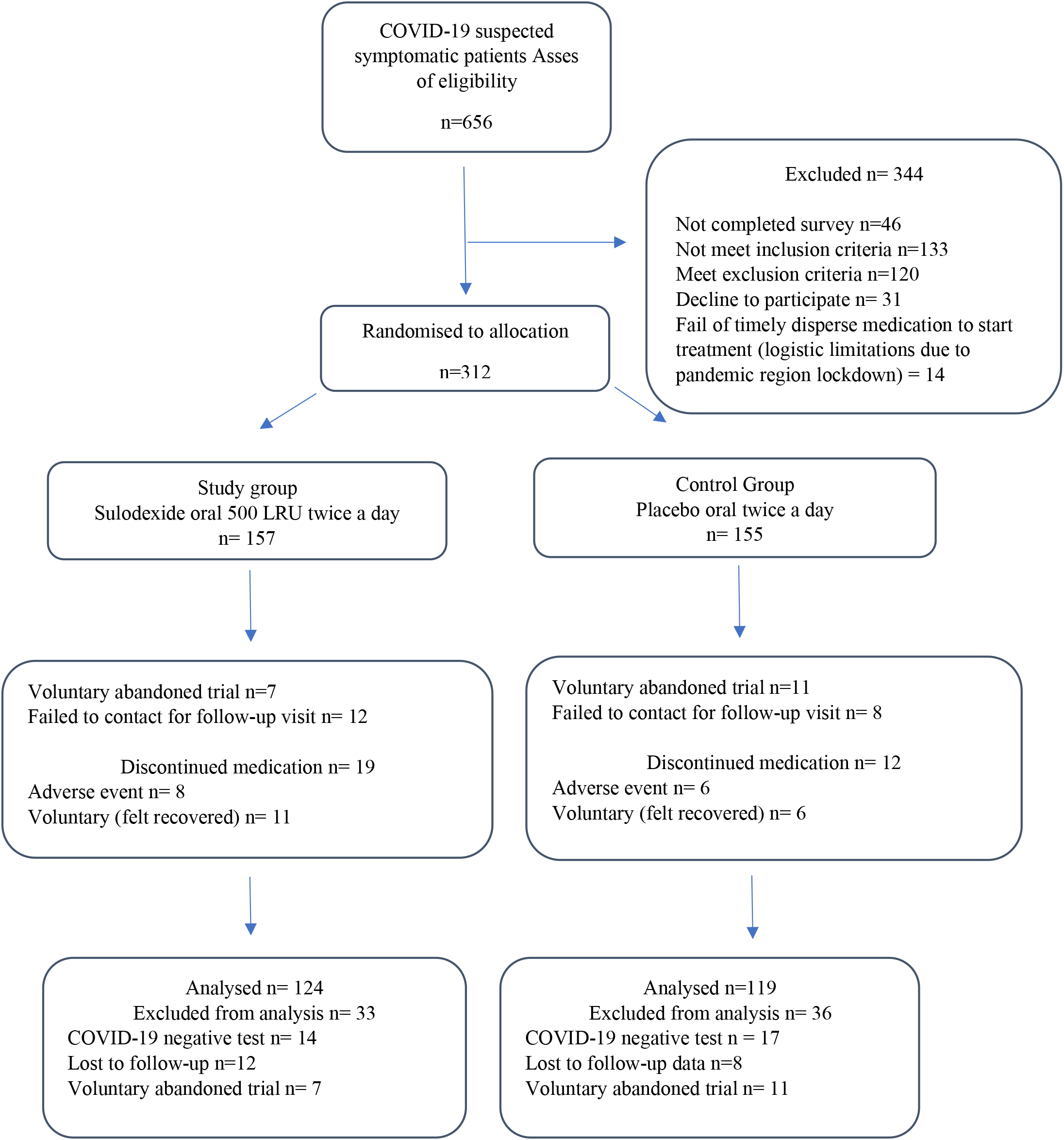
CONSORT flow diagram. Patients who discontinued medication were not excluded from the final analysis.

### Data sources

Data was collected using the Castor EDC software for validation and monitoring, and a hard copy file was maintained at the research site.

Data collected from each patient included the following: (1) patient general demographics; (2) clinical characteristics and outcomes; (3) serum test results as well as COVID-19 PCR test results; and (4) the duration and dosages of all therapies the participant received, adverse events and medication adherence.

### Study endpoints

Outcomes were assessed at 21 days post-randomisation. Primary endpoints were demarcated as the need for hospital admission and the total length of days (LOD) due to COVID-19 infection, need for oxygen support, and total days duration (including at home and hospital). Secondary endpoints were identified as follows: the need for mechanical ventilation support, presence of a thromboembolic event, major bleeding events, mortality, serum levels of D-dimer, C-reactive protein (CRP) and serum creatinine (Cr) levels.

### Statistical analysis

The need for hospital care was the study endpoint used to determine the sample size needed for statistical significance. Since we included a high-risk population, we anticipated a COVID-19 clinical progression that would warrant the need for hospital care of 40%. Using a t-test difference between two independent means with an effect size of 0·4, an alpha error of 0·05 and 80% power, we calculated a sample size of 100 subjects in each group. Planning a 20% incidence of attrition, we increased the sample size to 120 participants per group.

Estimates of risk ratios are presented with 95% Cis, and were calculated using the MedCalc software (MedCalc Software Ltd., Olsted,Belgium). All p-values are two-sided and are shown without adjustment for multiple testing. The complete database is maintained by the study team.

We express quantitative variables as mean (SD) and qualitative variables as frequencies and percentages. Differences in means were assessed using the Student’s t-test, while differences in percentages were assessed using the χ^2^ test. Before and after serum levels within the same patients were analysed using two paired t-tests. If data was not normally distributed, a non-parametric test was used

Although initially an intention-to-treat analysis had been proposed, the inclusion of only confirmed COVID-19 patients and the exclusion of patients who had initiated the treatment but later reported a negative COVID-19 result led to us analysing variables according to a per-protocol principle. The SPSS software (IBM SPSS Statistics for Windows, version 26, IBM Corp., Armonk, NY, USA) was used for data analysis. Missing data was present in less than 10% of the patients, and accounted for via multiple imputation analyses. An independent committee had access to safety data analysis.

## Results

Of the 312 patients randomised for group allocation, 31 patients reported a negative COVID-19 PCR test result [14 of 157 (8·9%) in the sulodexide group and 17 of 155 (10·9%) in the control group], and 38 patients had no sufficient data for analysis (voluntarily abandoned study or were lost to follow-up) [19 o 157 (12·1%) in the sulodexide group and 19 of 155 (12·2%) in the control group]. A total of 243 patients were eligible for final data analysis, 124 patients in the sulodexide group and 119 in the placebo group. The demographics, clinical characteristics and number of medications used by the patients were similar in both the groups. The median age was 52 ±10·6 years. Women accounted for 52·6% of the participants (128 of 243). Hypertension was the most common chronic health condition, reported in 34·2% (83 of 143), followed by diabetes 22·2% (54 of 243). The patient risk reported by the C19HC calculator was similar between groups (67·8% ±14 for the sulodexide group and 65·8% ±14·1 for the placebo group; p=0·32) (*table 3*).

**Table 3.** Results of general demographics, comorbidities, and outcome. * Percentage is given by the COVID-19 Health Complication (C19HC) risk calculator (Gobierno de Mexico, IMSS). † Includes the total of days that patients needed oxygen support at home or the hospital. Some patients continued oxygen support at-home after hospital care or started oxygen at home and later required hospital care. LOD = length of days, SD = standard deviation, n = number patients, % = percentage, BMI = body mass index. COPD = Chronic obstructive pulmonary disease. CRP = C-reactive protein.

|  | **Sulodexide**<br>**n= 124** | **Placebo**<br>**n= 119** | **Relative Risk**<br>**(95%CI)** | **P-value** |
| --- | --- | --- | --- | --- |
| **Demographics** |  |  |  |  |
| Age in years, mean (SD) | 55.3 (10.3) | 54 (10.9) | .. | 0.26 |
| Gender, n (%) |  |  |  |  |
| Male | 60 (48.3) | 55 (46.2) | 1.04 (0.80 - 1.36) | 0.73 |
| Female | 64 (51.6) | 64 (53.8) | 0.95 (0.75 - 1.21) | 0.73 |
| BMI, Mean (SD) | 29 (4.0) | 28.7 (3.2) | .. | 0.30 |
| **Chronic comorbidities, n (%)** |  |  |  |  |
| Diabetes mellitus | 22 (17.7) | 28 (23.5) | 0.75 (0.45 - 1.24) | 0.26 |
| Hypertension | 48 (38.7) | 35 (29.1) | 1.31 (0.92 - 1.87) | 0.13 |
| COPD | 30 (24.1) | 26 (21.8) | 1.10 (0.69 - 1.75) | 0.66 |
| Cardiovascular disease | 28 (22.5) | 23 (19.32) | 1.16 (0.71 - 1.90) | 0.53 |
| *C19HC risk calculator, mean (SD) | 67.87 (14.0) | 65.81 (14.1) | .. | 0.32 |
| **Outcome measures** |  |  |  |  |
| Need Hospital care, n (%) | 22 (17.7) | 35 (29.4) | 0.60 (0.37 - 0.96) | 0.03 |
| LOD hospital care, mean (SD) | 6.29 (4.1) | 7.8 (4.5) | .. | 0.21 |
| ‡Need Oxygen support, n (%) | 37 (29.8) | 50 (42.0) | 0.71 (0.50 - 1.00) | 0.05 |
| ‡LOD Oxygen support, mean (SD) | 9 (7.2) | 11.5 (9.6) | .. | 0.02 |
| Mortality | 3 (2.4) | 7 (5.8) | 0.41 (0.10 - 1.55) | 0.19 |
| Invasive Mechanical ventilation support, n (%) | 3 (2.4) | 6 (5.0) | 0.47 (0.12 - 1.87) | 0.29 |
| Haemodialysis, n (%) | 0 | 0 | .. | .. |
| Thromboembolic events, n (%) | 2 (1.6) | 2 (1.6) | 0.95 (0.13 - 6.70) | 0.96 |
| **Laboratory findings** |  |  |  |  |
| D-dimer, ng/dl |  |  |  |  |
| Baseline, mean (SD) | 293.6 (117.5) | 318.4 (131.6) | .. | 0.12 |
| < 500, n (%) | 110 (88.7) | 98 (75.6) | 1.07 (0.97 - 1.19) | 0.16 |
| > 500, n (%) | 14 (11.2) | 21 (17.6) | 0.63 (0.34 - 1.19) | 0.16 |
| Week 2, mean (SD) | 464.75 (629.81) | 897.76 (1215.36) | .. | <0.01 |
| < 500, n (%) | 97 (78.3) | 63 (52.95) | 1.47 (1.21 - 1.79) | <0.01 |
| > 500, n (%) | 27 (21.7) | 56 (47.05) | 0.46 (0.31 - 0.67) | <0.01 |
| CRP, mg/dl |  |  |  |  |
| Baseline, mean (SD) | 10.6 (6.4) | 10.1 (6.9) | .. | 0.55 |
| Week 2, mean (SD) | 12.55 (10.2) | 17.81 (11.56) | .. | <0.01 |
| Creatinine week 2, mg/dl |  |  |  |  |
| < 1.6 | 113 (91.1) | 107 (89.9) | 1.01 (0.93 - 1.09) | 0.74 |
| > 1.6 | 11 (08.8) | 12 (10.0) | 0.87 (0.40 - 1.91) | 0.74 |

### Primary outcome

Overall, 57 of 243 patients (23·4%) required hospital care during the 21 days of follow-up [22 of 124 (17·7%) in the sulodexide group and 35 of 119 (29·4%) in the placebo group with a relative risk (RR) of 0·6; 95% confidence interval (CI) 0·37 to 0·96; p=0·03]; a number needed to treat (NNT) for benefit of 8·5. The mean hospital LOD was 6·2 ±4·1 in the sulodexide group vs. 7·8 ±4·5 in the placebo group; p=0·21(Figure 2). There were 87 of 243 patients (35·8%) who developed respiratory symptoms requiring oxygen support [37 of 124 (29·8%) in the sulodexide group vs. 50 of 119 (42%) in the control group with an RR of 0·71; 95% CI 0·5 to 1; p=0·05]. The mean LOD of oxygen support was 9 ±7·2 in the sulodexide group vs. 11·5 ±9·6 in the placebo group; p=0·02 (Table 3).

**Figure 2.**
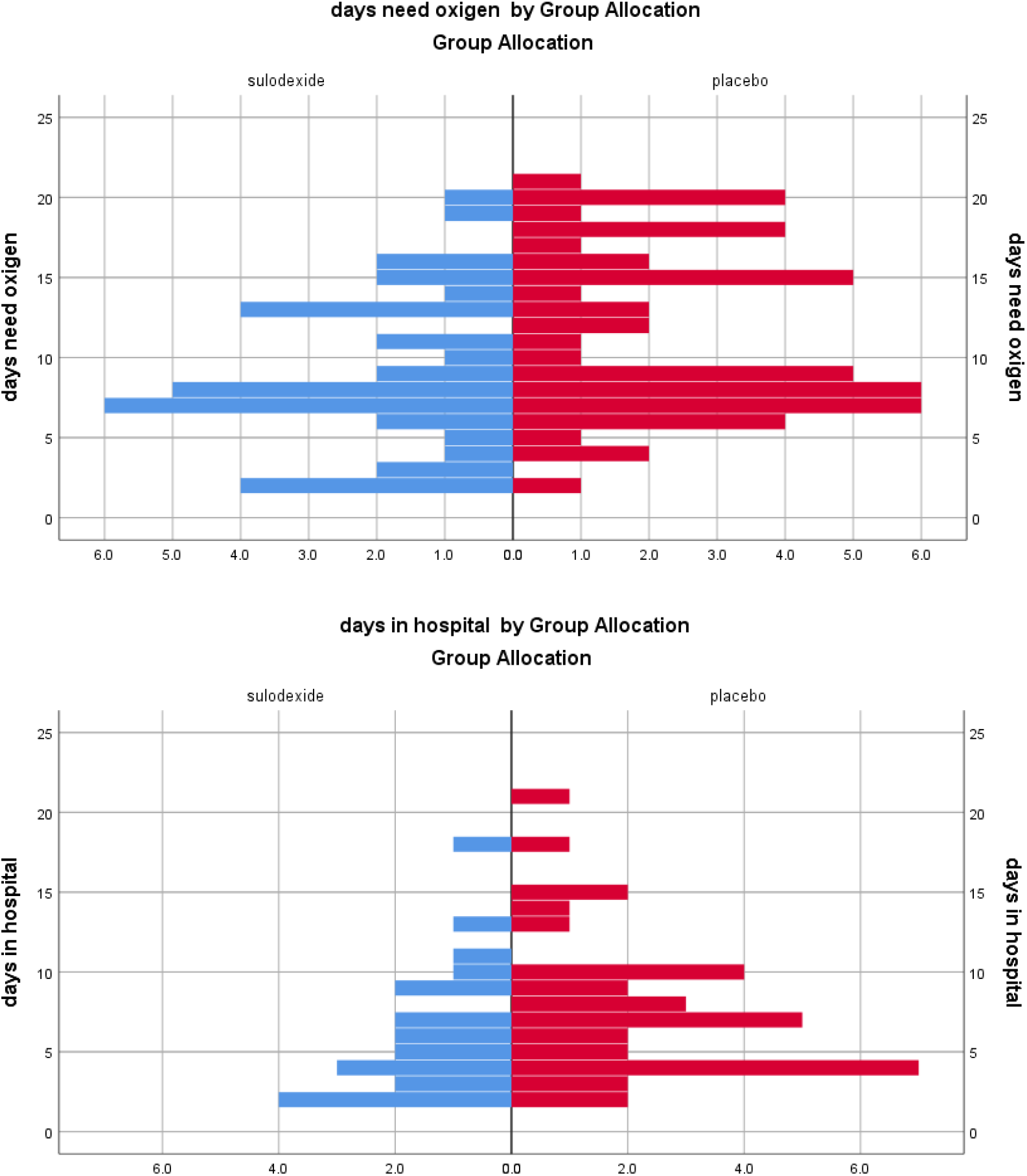
Population pyramid frequency. Total days of hospital stay and for the need for oxygen support. It is not showing patients with 0 days.

### Secondary endpoints

Importantly, the overall mortality rate was 4·1% (10 of 243), comprising of 2·4% (3 of 124) in the sulodexide group vs. 5·8% (7 of 119) in the placebo group; RR of 0·41; 95% CI 0·10 to 1·55; p=0·19 (*figure 3*). The mean D-dimer levels at week 2 were significantly elevated in the placebo group compared to the sulodexide group (897·7 ±1215·36 vs. 464·75 ±629·81; p<0·01). There were 27 of 124 (21·7%) patients in the sulodexide group who showed a D-dimer value >500 ng/dl vs. 56 of 119 (47·05%) in the placebo group with a RR of 0·46; 95% CI 0·31 to 0·67; p>0·01. The mean C-reactive protein level at week 2 was 12·55 ±10·2 mg/dL in the sulodexide group vs. 17·81 ±11·56 mg/dL in the placebo group; p<0·01 (*Figure 4*).

**Figure 3.**
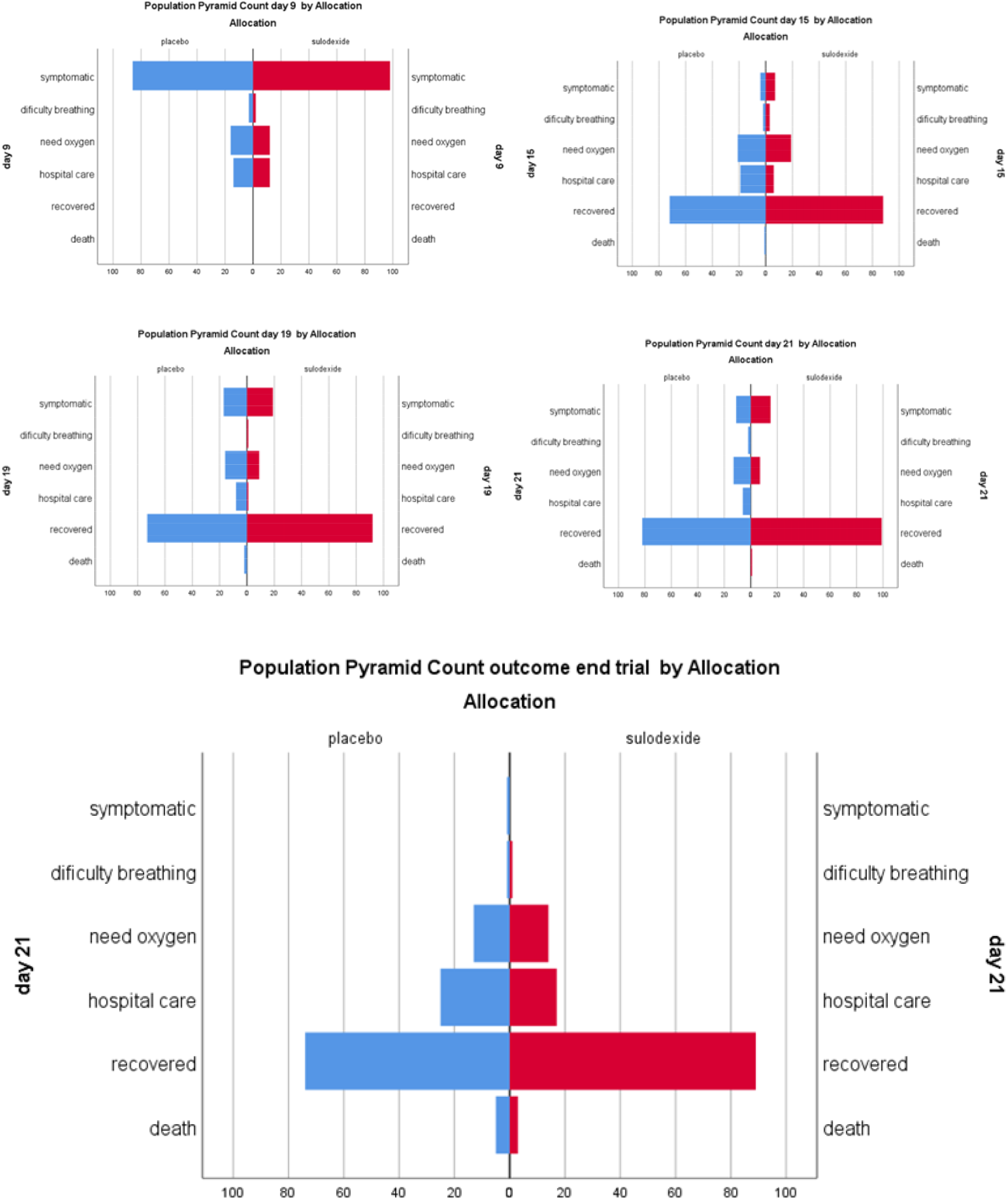
Outcome progression. Population pyramid count of outcome during the follow-up period. For this chart, recovered was considered if symptoms were mild enough to permit regular activities or return to work. Some patients that needed oxygen support are included in the hospital care cluster. The graph includes two patients whose death occurred after the 21 days follow-up.

**Figure 4.**
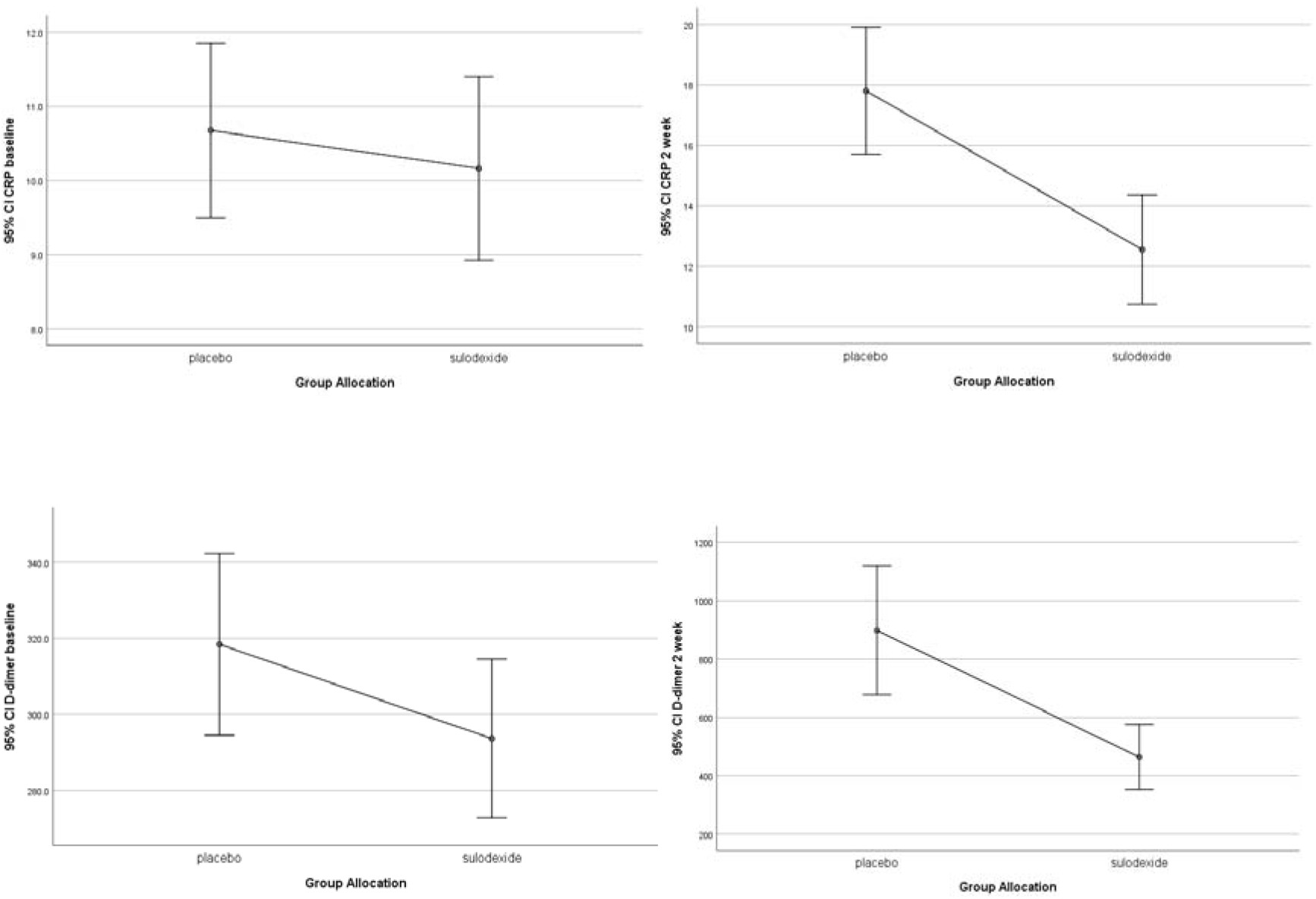
Laboratory results. Mean D-dimer and C-reactive protein (CRP) serum levels according to group allocation at baseline and two weeks.

### Adherence and safety

Medication adherence was evaluated via the questionnaire conducted at every follow-up visit, along with indirect verification by counting the residual capsules in the medication blisters. A total of 19 of 124 (15·3%) in the sulodexide group vs. 12 of 119 (10%) in the control group suspended their medication with a RR of 1·81; 95% CI 0·88 to 3·74; p= 0·10. A total of 17 of 243 (13·7%) of the patients felt clinically recovered, resulting in voluntary premature interruption of medication 11 of 124 (8·8%) in the sulodexide group vs. 6 of 119 (5%) in the control group for a RR of 0·56; 95% CI 0·21 to 1·48; p= 0·24 between group difference. This interruption occurred after a minimum of 14 days of treatment in all the patients, none of whom required later hospital care or oxygen support. None of the patients was excluded from the final analysis.

Novel symptoms reported by the patients after initiating sulodexide or placebo intake were also reported, although many could have been a delayed onset of COVID-19 symptoms. A new event was present in 181 of 243 patients (74·4%) with an RR of 1·08; 95% CI 0·93 to 1·25; p=0·28 between-groups comparison. An adverse event was severe enough to cause medication cessation in 14 of 243 patients (5·7%) with an RR of 0·78; 95% CI 0·27 to 2·18; p=0·63 between-groups comparison; gastrointestinal discomfort was stated as the main reason for suspension. Only one patient (part of the control group) presented a major bleeding event (define as bleeding causing a fall in haemoglobin level of 2 g/dL or more, or leading to transfusion of two or more units of whole blood or red cells). This event occurred while on hospital care, and eventually was fatal. The full list of adverse events has been provided in *table 4*.

**Table 4.** Medication adherence and adverse events. Values are through day-21, the date of the scheduled completion of the trial intervention. † Patients while on the per-protocol outpatient setting. The main reason for the voluntary suspension of medication was symptoms improvement. * Three patients in the control group and five patients in the study group that suspended medication due to an adverse event required hospital care due to severe clinical disease progression. ** No patient that suspended medication for voluntary reasons needed hospital care or oxygen support. ‡More than one adverse event could occur per patient. CI= confidence interval.

|  | **Sulodexide**<br>**N=124** | **Placebo**<br>**N=119** | **Relative risk**<br>**(CI 95%)** | **P-value** |
| --- | --- | --- | --- | --- |
| **Medication adherence, n, (%)** |  |  |  |  |
| †All the time | 91 (73.3) | 99 (83.1) | 1.13 (0.99 - 1.29) | 0.06 |
| †Most of the time | 14 (11.2) | 8 (6.7) | 0.59 (0.25 - 1.36) | 0.22 |
| Suspended medication |  |  |  |  |
| total | 19 (15.3) | 12 (10) | 1.81 (0.88 - 3.74) | 0.10 |
| *Adverse event | 8 (6.4) | 6 (5) | 0.78 (0.27 - 2.18) | 0.63 |
| ** Voluntary | 11 (8.8) | 6 (5) | 0.56 (0.21 - 1.48) | 0.24 |
| **Adverse event, n, (%)** |  |  |  |  |
| ‡Total | 96 (77.4) | 85 (71.4) | 1.08 (0.93 - 1.25) | 0.28 |
| Abdominal discomfort (gastritis, nausea, vomiting or diarrhoea). | 36 (29) | 39 (32.7) | 1.12 (0.77 - 1.64) | 0.52 |
| Headache. | 96 (77.4) | 85 (71.4) | 0.92 (0.79 - 1.07) | 0.28 |
| Major Bleeding. | 0 | 1 (0.8) | 3.12 (0.12 - 75.96) | 0.48 |
| Skin reaction. | 3 (2.41) | 5 (4.2) | 1.73 (0.42 - 7.10) | 0.44 |

The outpatient treatment recommendations in the study population were heterogeneous, but evenly distributed between groups. Of importance, the use of inhaled bronchodilators was lower in the sulodexide group [70 of 124 (56·45) patients vs. 85 of 119 (71·42) with a RR 0·79; 95% CI 0·65 to 0·95; p=0·01]. A list of concomitant medications is reported in *table 5*.

**Table 5.** Concomitant medication. The list includes only the medications used in the outpatient setting. Patients usually received more than one additional medication. NSAID= Non-steroidal anti-inflammatory drugs. NOAC = Non-Vitamin K antagonist oral anticoagulants (novel oral anticoagulants). LMWH = Low molecular weight heparin. ACE = Angiotensin-converting enzyme.

|  | **Sulodexide**<br>**Group**<br>**n=124** | **Placebo**<br>**Group**<br>**n=119** | **RR**<br>**(95% CI)** | **p-value** |
| --- | --- | --- | --- | --- |
| **Medication before trial, n, (%)** |  |  |  |  |
| Aspirin | 33 (26·61) | 44 (36·97) | 0·71 (0·49 – 1·04) | 0·08 |
| Oral hypoglycaemics | 19 (15·32) | 27 (22·68) | 0·67 (0·39 – 1·14) | 0·14 |
| Insulin | 14 (11·29) | 18 (15·12) | 0·74 (0·38 – 1·43) | 0·37 |
| ACE inhibitors | 22 (17·74) | 32 (26·89) | 0·65 (0·40 – 1·06) | 0·09 |
| Other anti-hypertensive | 45 (36·29) | 41 (34·45) | 1·05 (0·74 – 1·48) | 0·76 |
| Statins | 14 (11·29) | 16 (13·44) | 0·83 (0·42 – 1·64) | 0·61 |
| **Medication added during trial, n, (%)** |  |  |  |  |
| Paracetamol | 78 (62·90) | 82 (68·90) | 0·91 (0·76 – 1·09) | 0·32 |
| LMWH | 12 (9·67) | 16 (13·44) | 0·71 (0·35 – 1·45) | 0·36 |
| NOAC's | 9 (7·25) | 11 (9·24) | 0·78 (0·33 – 1·82) | 0·57 |
| Ivermectin | 54 (43·54) | 59 (49·57) | 0·87 (0·67 – 1·15) | 0·34 |
| Hydroxychloroquine | 46 (37·09) | 36 (30·25) | 1·22 (0·85 – 1·75) | 0·26 |
| Corticoids | 79 (63·70) | 73 (61·34) | 1·17 (0·97 – 1·40) | 0·08 |
| Statins | 25 (20·16) | 21 (17·64) | 1·14 (0·67 – 1·92) | 0·61 |
| Vitamins – supplements | 95 (76·61) | 101 (84·87) | 0·90 (0·79 – 1·02) | 0·10 |
| Antibiotics | 41 (33·06) | 35 (29·41) | 1·12 (0·77 – 1·63) | 0·54 |
| Other NSAID | 67 (54·03) | 56 (47·05) | 1·14 (0·89 – 1·47) | 0·27 |
| Omeprazole | 98 (79·03) | 105 (88·23) | 0·88 (0·78 – 0·98) | 0·03 |
| Antacids | 44 (35·48) | 36 (30·25) | 1·17 (0·81 – 1·68) | 0·38 |
| Inhaled bronchodilators | 70 (56·45) | 85 (71·42) | 0·79 (0·65 – 0·95) | 0·01 |
| Oseltamivir | 33 (26·61) | 28 (23·52) | 1·13 (0·73 – 1·74) | 0·58 |

## Discussion

This study evaluated the therapeutic effect of sulodexide in patients with early stages of COVID-19 in a real-life setting. Sulodexide was effective in decreasing the need for hospital admission and oxygen requirements. In addition, lower serum levels of inflammatory and prothrombotic markers were also observed. Our findings support the effectiveness of sulodexide in mitigating the severe clinical progression rate of COVID-19, compared to the prevalent standard of care, when used in the early symptomatic stages of the disease.

The clinical progression of COVID-19 shows a biphasic pattern. The first phase corresponds to virus replication characterised by upper respiratory symptoms, following which ∼80% of the patients begin recovery. The second phase is associated with a severe inflammatory response and is characterised by symptom persistence, the onset of mild breathing difficulty and chest pain. These symptoms can rapidly progress to full acute respiratory distress syndrome (ARDS), which necessitates supplemental oxygen or hospital care^25,26^. The study endpoints of the trial focus on this stage of the disease. The use of the C19HC calculator helped identify patients at higher risk of progressing to this phase by clustering age and different chronic comorbidities into a numeric risk value^21^. Thus, the need for a large sample size to achieve primary outcome significance was successfully overcome, along with the related financial and logistic implications. Although the need for oxygen support is the most common indication for hospital admission in COVID-19 patients^27^, if the oxygen supply can be allocated at home and no other specific medication or care is needed, these patients can continue treatment at home. For this reason, supplemental oxygen was included alongside hospital care as a disease severity progression endpoint.

An important finding was that almost 15% of patients already showed elevated levels of D-dimer at baseline, highlighting the importance of early action. Although sulodexide might be better known for its antithrombotic effect— similar to that of low molecular weight heparin (LMWH)— and has been proposed as an option for targeting thromboembolic risk in COVID-19 patients^28^, its endothelial protective properties ^29,30^ may add a benefit of equal or greater importance in the early stages of the disease. Sulodexide can be used with no significant risk of side effects, which limits the widespread use of LMWH and corticoids early on.

The high incidence of adverse events reported resulted from the difficulty in establishing which events were specific to sulodexide, since some symptoms could be related to COVID-19 or one of the many medications administered to these patients. Headache and gastrointestinal discomfort were the most common adverse effects, which could be attributed to the high level of stress in the treatment population due to prolonged confinement and uncertainty of disease progression.

Although the number of confirmed thromboembolic events was low and similar in both the groups, we could not properly verify this outcome. Poor access to ultrasound or computer tomography limited the verification in some patients with high clinical suspicion. Moreover, the D-dimer levels could potentially be an indirect marker representing the prevention benefit of sulodexide against these events. We observed that once the patient required hospital care, there was no between-group difference with respect to total days of hospital care, the need for mechanical ventilation or haemodialysis. These findings suggest that once the clinical setting is critical, there can be an overlay of complications related to severe systemic disease.

In the initial protocol design, mortality was intended to be a primary endpoint. However, final mortality data resulted underpowered for relevant analysis, and was reported as a secondary endpoint instead. Mortality was numerically lower in the sulodexide group, but the difference was not statistically significant. The difference between the 4·1% mortality rate seen in this trial vs. the national average of 10·4% in Mexico^3^ can be explained by the number of participants tested, since to date, Mexico has not initiated widespread testing.

This trial does pose certain acknowledgeable limitations. Numerous asymptomatic carriers and lack of widespread diagnostic testing worldwide make it difficult to establish a real incidence of the disease, affecting accurate sample size calculations. With a significant part of the trial taking place in an outpatient setting, we did not foresee some of the logistic and infrastructure limitations due to the region’s pandemic lockdown. Key members of the staff were unavailable during part of the trail which forced the lead researcher to undertake data managing duties, breaking his blinding to group allocations. Other restrictions were also experienced in the hospital setting, for example, the impossibility to perform post-mortem examination. The different numbers and types of medications prescribed to the study population were very heterogenic. Moreover, there existed a noticeable difference between the prescription from primary physicians in the private sector vs. government care centres. Although evenly distributed among groups, the above factors should be taken into consideration during the interpretation of the study results.

There is also reasoning for a later exploratory trial from an economic viewpoint. A significant benefit could be found when considering the total number of patient-hospital days (number of patients × mean number of hospital LOD) of 136 in the sulodexide group vs. 273 in the placebo group, with the consumed resources and cost 1-patient/day can represent. In addition, sulodexide is less expensive in the Mexican market than LMWH or other new oral anticoagulants (NOACs).

In summary, when used in the early stages of COVID-19, the synergic activity of sulodexide’s pleiotropic effects on different biological targets can play an essential role in limiting disease progression, resulting in a decreased need for oxygen support and hospital care, as observed in this trial. This has beneficial implications in both patient well-being and economics, potentially making sulodexide a favourable medication until an effective vaccine or an antiviral becomes available. These findings justify further multicentre confirmatory studies.

This trial is listed in the ISRCTN registry under the study ID ISRCTN59048638.

## Data Availability

The data analyzed and presented in this study are available from the corresponding author on reasonable request, providing the request meets local ethical and research governance criteria.

## Data sharing

The data analysed and presented in this study are available from the corresponding author on reasonable request, providing the request meets local ethical and research governance criteria.

## References

[1] World health organisation, “COVID-19 Early Epidemiologic and Clinical investigations for public health,” February 2020. [Online]. Available: https://www.who.int/docs/default-source/coronaviruse/200218-early-investigations-one-pager-v1-eng.pdf?sfvrsn=8aa0856_14. [Accessed 15 April 2020].

[2] F. Dawood, P. Ricks, B. Njie, M. Daugherty, W. Davis, J. Fuller and e. al, “Observations of the global epidemiology of COVID-19 from the prepandemic period using web based surveillance: a cross sectional analysis,” Lancet Infect Dis, July 29 2020.

[3] Gobierno de Mexico, “Secretaria de Salud,” 07 Jun 2020. [Online]. Available: https://coronavirus.gob.mx/datos/. [Accessed 07 Jun 2020].

[4] M. Vaduganathan, O. Vardeny, D. Pharm, T. Michel, J. McMurray, M. Pfeffer and S. Solomon, “Renin– Angiotensin–Aldosterone System Inhibitors in Patients with Covid-19,” N Engl J Med, vol. 382, no. 17, pp. 1653–1659, April 23 2020.

[5] M. Ratajczak, K. Bujko, A. Ciechanowicz, K. Sielatycka, M. Cymer, W. Marlicz and e. al., “SARS-CoV-2 Entry Receptor ACE2 Is Expressed on Very Small CD45-Precursors of Hematopoietic and Endothelial Cells and in Response to Virus Spike Protein Activates the Nlrp3 Inflammasome,” Stem Cell Rev and Rep, pp. 1–12, July 20 2020.

[6] L. Carsana, A. Sonzogni, A. Nars, R. Rossi, P. A, P. Zerbi and e. al., “Pulmonary post-mortem findings in a series of COVID-19 cases from northen Italy: a two-centre descriptive study,” Lancet Infect Dis, pp. 1–6, June 8 2020.

[7] A. Schutte and D. Harrison, “Immunity, inflammation and the vasculature in the COVID-19 era,” Journal of hypertension, vol. 38, no. 9, pp. 1701–1702, Sep 2020.

[8] J. Moore and C. June, “Cytokine release syndrome in severe COVID-19,” Science, vol. 368, no. 6490, pp. 473–474, May 1 2020.

[9] M. Ackermann, S. Verleden, M. Kuehnel, A. Haverich, T. Welte, F. Laenger and e. al., “Pulmonary Vascular Endothelialitis, Thrombosis, and Angiogenesis in Covid-19,” NEJM, May 21 2020.

[10] B. Becker, M. Jacob, S. Leipert, A. Salmon and D. Chappell, “Degradation of theendothelial glycocalyx inclinical settings: searching forthe sheddases,” Br J Clin Pharmacol, vol. 80, no. 3, pp. 389–402, March 16 2015.

[11] P. Evans, G. Rainger, M. J, T. Guzik, E. Osto, Z. Stamataki and e. al., “Endothelial dysfunction in COVID-19: a position paper of the ESC Working Group for Atherosclerosis and Vascular Biology, and the ESC Council of Basic Cardiovascular Science,” Cardiovascular Research, Aug 2020.

[12] R. Cao, L. Tang, Z. Xia and R. Xia, “Endothelial glycocalyx as a potential theriapeutic target in organ injuries,” Chin Med J, pp. 963–975, April 20 2019.

[13] S. Coccheri and F. Mannello, “Development and use of sulodexide in vascular diseases: implications for treatment,” Drug Des Devel Ther, vol. 8, pp. 49–65, 24 Dic 2013.

[14] T. Li, X. Liu, Z. Zhao, L. Ni and C. Liu, “Sulodexide recovers endothelial function through reconstructing glycocalyx in the balloon-injury rat carotid artery model,” Oncotarget, vol. 8, no. 53, pp. 91350–91361, October 31 2017.

[15] A. Zielinski, M. Zabel, T. Wysocka, T. Urbanek and K. Suminska, “Sulodexide activates glycocalyx restorations in patients with chronic venous disease,” Vascular Insight Nautilus, pp. 17–18, 2019.

[16] V. Masola, G. Zaza, M. Onisto, A. Lupo and G. Gambaro, “Glycosaminoglycans, proteoglycans and sulodexide and the endothelium: biological roles and pharmacological effects,” Int Angiol, vol. 3, no. 33, pp. 243–254, Jun 2014.

[17] F. Mannello, D. Ligi, M. canale and J. Raffetto, “Sulodexide Down-Regulates the Release of Cytokines, Chemokines, and Leukocyte Colony Stimulating Factors from Human Macrophages: Role of Glycosaminoglycans in Inflammatory Pathways of Chronic Venous,” Current Vascular Pharmacology, vol. 12, no. 1, pp. 173–185, 2014.

[18] P. Matta, F. Manello, P. Ferrari and G. Augus, “Vascular pathologies and inflammation: The anti-inflammatory properties of sulodexide,” Italian Journal of Vascular endovascular surgery, pp. 1–7, January 2012.

[19] G. Pompilio, D. Integlia, J. Raffetto and G. Palareti, “Comparative Efficacy and Safety of Sulodexide and Other Extended Anticoagulation Treatments for Prevention of Recurrent Venous Thromboembolism: A Bayesian Network Meta-analysis,” TH open, vol. 4, no. 2, pp. E80–E93, April 28 2020.

[20] gobierno de Mexico, “Recomendaciones para el tratamiento de la infeccion por SARS-CoV-2, agete causal de COVID-19,” 06 Jul 2020. [Online]. Available: https://coronavirus.gob.mx/wp-content/uploads/2020/07/Recomendaciones_para_tratamiento_SARS-CoV2.pdf. [Accessed 20 Aug 2020].

[21] Gobierno de Mexico: IMSS, “Calculadora de complicacion de salud por COVID-19,” 05 2020. [Online]. Available: http://www.imss.gob.mx/covid-19/calculadora-complicaciones. [Accessed 06 06 2020].

[22] G. Andreozzi, A. Bignamini and G. Davi, “Sulodexide for the prevention of recurrent venous thromboembolism: The SURVET study: a Multicenter, Randomised, double-blind, placebo-controlled trial,” Circulation, vol. 132, no. 20, pp. 1891–1897, 2015.

[23] gobierno de mexico, secretaria de Salud, lineamientos para la atencion de pacientes con COVID-19, Ciudad de Mexico, 2020.

[24] Instituto Mexicano Seguro Social, “Algoritmos interinos para la atencion del COVID-19. Actualizacion 31 de Julio del 2020,” 31 July 2020. [Online]. Available: http://educacionensalud.imss.gob.mx/es/system/files/Algoritmos_interinos_COVID19_CTEC.pdf. [Accessed 2020].

[25] S. Richardson, J. Hirsch, M. Narasimhan, J. Crawford, T. McGinn, K. Davidson and e. al., “Presenting Characteristics, Comorbidities, and Outcomes Among 5700 Patients Hospitalised With COVID-19 in the New York City Area,” JAMA, pp. 2052-2059, April 22 2020.

[26] J. Wise, “Covid-19: Study reveals six clusters of symptoms that could be used as a clinical prediction tool,” BMJ, pp. 1–2, July 20 2020.

[27] J. Chen, T. Qi, L. Liu, Y. Ling, Z. Qian, T. Li and e. al., “Clinical progression of patients with COVID-19 in Shanghai, China,” Journal of infection, vol. 80, no. 5, pp. e1–e6, May 2020.

[28] B. Bikdeli, M. Madhavan, A. Gupta, D. Jimenez, J. Burton, C. Nigoghossian, T. Chuich and e. al., “Pharmacological Agents Targeting thomboinflammation in COVID-19:review and implications for future research,” Thromb haemost, vol. 120, no. 7, pp. 1004–1024, May 20 2020.

[29] T. Urbanek, K. Zbigniew, B. Begier-Krasińska, E. Baum and A. Bręborowicz, “Sulodexide suppresses inflammation in patients with chronic venous insufficiency,” Int Angio, vol. 35, no. 2, pp. 140–147, december 2016.

[30] A. Połubińska, R. Stanisewisky, E. Baum, K. Suminska and A. Brevorowicz, “Sulodexide modifies intravascular homeostasis what affects function of the endothelium,” advances in medical sciences, vol. 58, no. 2, pp. 304–310, 2013.

